# Assessment of compliance of baby friendly hospital initiative in tertiary care hospital, South India

**DOI:** 10.1101/2023.07.14.23292661

**Authors:** A.charles pon ruban, Lekshmy S Nair, Linta Maria David, Mahalakshmi V, Manorita Jerin Grace, Manoj Prithvi K, Jyodhish K S

## Abstract

**Objectives:** To assess the compliance of a tertiary care hospital, South India with the ten steps of the Baby Friendly Hospital Initiative following the UNICEF/WHO global criteria.

**Design:** Cross-sectional study

**Setting:** Tertiary care hospital, South India

**Participants:** Staff nurses who are posted in in the obstetrics and gynaecology department and the mothers admitted antenatal, postnatal, and labour wards.

**Results:** Only 35% of the staff nurses interviewed received formal training in breastfeeding techniques. Written information about breastfeeding was given to all prenatal mothers (100%) yet, only 75% knew the importance of breastfeeding soon after delivery. Only 62.5% of babies were exclusively breastfed and 51.3% of the babies were breastfed within one hour of birth. Compliance with the Ten Steps of baby friendly initiative was 66.63%, signifying a moderate compliance overall.

**Conclusion:** The compliance to Ten steps of BFHI in low resource setting shows an overall moderate compliance. The Ten Steps of BFHI may be implemented more effectively in all medical facilities through the mother’s absolute affection Programme (MAA). More focused approaches are needed to improve the breastfeeding practices even in tertiary care settings in India.

**Keypoint:** *What is already known on this topic:* Good breast-feeding practices are essential for reducing infant mortality. The effect of interventions by the Government on breast feeding practices especially in a tertiary care hospital setting, South India is poorly studied.

*What this study adds:* Practice of exclusive breast feeding is low. Compliance with the Ten Steps of baby friendly initiative was low even in a tertiary care setup in south India.

*How this study might affect research, practice and/or policy:* Good breast-feeding practices should be promoted in all health facilities. More focused interventions are needed to fill this gap.

## INTRODUCTION

Breastfeeding has been recognized as an essential component of a child’s nutrition because of its dramatic effects on morbidity and mortality, especially in infants under one year of age. According to estimates, achieving universal breastfeeding rates might stop 823,000 children from dying each year.^[1]^Malnutrition is directly or indirectly responsible for more than two thirds of the 10.9 million deaths of children under five worldwide each year. More than two thirds of infant deaths under the age of five occur in the first year of life, and inappropriate feeding practices are frequently to blame. In accordance with recommendations from the World Health Organization (WHO) and the United Nations International Children’s Emergency Fund (UNICEF), breastfeeding should begin within an hour of delivery, be continued exclusively for the first six months of a child’s life and continued for at least two more years with safe and suitable complementary foods. According to global data on newborns and children, only 40% of infants under the age of six months are exclusively breastfed, and only 44% of infants begin nursing within the first hour of life. Only 45% of children are breastfed until 2 years of age. ^[2]^ The risk of death in the first 28 days of life is 33% higher for infants who begin breastfeeding between two and twenty-three hours after birth, and more than twice as high for those who begin one day or longer after birth, compared to newborns who were placed to the breast within the first hour after birth.^[3]^

The WHO and UNICEF’s collaborative programme, the Baby Friendly Hospital Initiative (BFHI), has identified 10 actions to support, safeguard, and institutionalize breastfeeding in maternity and paediatric hospitals around the world. ^[4]^ The Ten Steps can be thought of as a quality assessment and improvement system because they are based on specific actions in five breastfeeding areas. (1) Policy: BF policies should support the 1981 WHO Code and the Ten Steps, and they should forbid the direct or indirect marketing of infant formula to women. (2) Training in BF for staff members of maternity wards. (3) Promotion and assistance: starting BF early in pregnancy, BF when needed, and community help, such as in-house support groups and referrals. (4) Prevention: Avoid using pacifiers or teats and avoid baby formula. (5) Restructuring the maternity ward workflow by requiring patients to stay in their rooms during their hospital stay. According to the Baby Friendly Hospital Initiative, the “Ten Steps to Successful Breastfeeding” should be the standard of care for all maternity care facilities in order to protect, encourage, and promote breastfeeding.^[5]^ Employees have the opportunity to promote BFHI practises and identify issues with implementation during reassessment. Being baby-friendly is a gradual process rather than something that happens all at once. The BFHI must be created as a multi-level, evidence-based change process that can identify effective strategies for greater inclusion in healthcare delivery.^[6]^ Mother’s absolute affection Programme(MAA) is a programme launched by Government of India (GOI) in 2016 to promote breastfeeding through health systems.^[7]^ According to the World Breastfeeding Trends Initiative (WBT)-India study from 2018, which highlighted that there has been no development since 2005 and that India’s implementation of the BFHI initiative is not performing well, there are significant deficiencies in both policy and programmes. ^[8,9]^.

There aren’t much research that have looked at the obstacles to the adoption of Baby-Friendly behaviors that may be used to boost BFI uptake at the local or national levels. In this setting, it becomes essential to assess the compliance of tertiary care hospital in a government sector with baby friendly hospital initiative.

## OBJECTIVES

To assess the compliance of a tertiary care hospital, South India with the ten steps of the Baby Friendly Hospital Initiative following the UNICEF/WHO global criteria.

## METHODS

### Study setting

Tertiary care hospital, Tamil Nadu, South India

### Study design

Cross sectional study

### Study duration

October 2022 to November 2022 (1 month)

### Sample size

Sample size was calculated using the following formula: 4 PQ/d2.

Prevalence of 86 % (BFHI compliance)^[10]^ and absolute precision of 5% were considered. The expected sample size was obtained to be 180, divided between Antenatal mother (n=80), Post natal mothers (n=80) and staff nurses (n=20)

### Sampling method

Simple random sampling

### Study population

Staff nurses who are posted in in the obstetrics and gynaecology department and the mothers admitted antenatal, postnatal, and labour wards.

- Eligibility criteria:
  □ Eligibility for the pregnant women was at least two ANC visits and the pregnancy in the third trimester.
  □ Eligibility for postpartum mothers was receiving ANC in the study facility, recording at least three ANC visits during the pregnancy, and delivering after 32 gestational weeks. Also, they should have been booked for discharge and received discharge counseling.
  □ Eligibility for the staff entailed caring for pregnant women, mothers, and babies for at least 6 months.

### Exclusion criteria

Postpartum mothers whose babies were unhealthy or preterm were excluded.

### Procedure

Compliance was measured using The UNICEF/WHO BFHI External Reassessment Tool developed in 2006 based on global Baby-Friendly key indicators.^[11]^ The assessment tool is subdivided into three main parts. The first and second parts are used for data collection, and the third part is used to transfer results, tally, and score. The structured questionnaires contain quantitative questions with categorical responses, and some open-ended qualitative probes, but no qualitative analysis is needed.

The reassessment tools are standardized global criteria developed after wide consultations and multi-country field testing and are proven to be applicable in all maternity settings regardless of the simplicity or sophistication (WHO & UNICEF, 2009). The questionnaire was translated into the local vernacular language and tested for face validity.

### Ethical committee approval

Ethical approval was obtained from the relevant Ethics Committee prior to the commencement of the study. All participants were provided with written information about the content and the purpose of the study. All participants gave their written consent to record the data. The participants’ confidentiality and autonomy were protected at all stages.

### Data collection

The Baby friendly hospital initiative monitoring and reassessment: tools to sustain progress, World health organization, Geneva data collection instrument was used.^[12,13]^

The participants were instructed to go through the questionnaires and get their doubts clarified before answering. For those unable to read, the information was disclosed in the presence of a witness and data collected.

#### Qualitative Data Collection

The first phase of the reassessment process involved assessing the maternal/newborn profile of the hospital, reviewing documents, and observing procedures in key areas. The document reviews and observations were scored based on adequacy, accuracy, and completeness. Possible scores ranged from zero to 100%.

#### Quantitative Data Collection

The second phase of the process involved face-to-face interviews to assess (a) knowledge (e.g., what is the most common cause of insufficient milk?); (b) skills (e.g., can you show me how you position your baby for breastfeeding?); (c) practices (e.g., how soon after birth was your baby given to you?); and (d) support systems (e.g., to whom do you refer mothers for help with milk expression?).

“Yes” responses were equivalent to 100 points; “No” and “Don’t know” to 0 points. Unanswered statements were assigned 0 points as the practice was considered compliant only if the respondent was aware of it.

### Data entry

Data entry was done in Microsoft Excel as per the standard BFHI monitoring and reassessment tool.

### Data analysis

Results were entered into the WHO/UNICEF BFHI computer tool (WHO & UNICEF, 2009), summarized, and scored. All data were analyzed quantitatively and presented as descriptive statistics. Percentage scores were awarded for the document reviews (“yes” and “no”), observation of areas (“yes”, “no,” and “area does not exist”), observation of procedures (number of correct observations out of the total observations), and interviews (number of correct responses out of the total interviews). In the case of document reviews and observation of areas, a “yes” (equivalent to 100%) was awarded when a policy was available and “no” (equivalent to 0%) when it was unavailable. Percentage scores for each criterion were summed, and the average was taken to estimate the total compliance. The global standards require a minimum of 80% compliance for almost all indicators; therefore, a cumulative average score above 80% signified a pass. To determine the extent of the BFHI implementation, compliance was classified as low (< 50%), moderate (50–80%), and high (> 80%).

### Patient and public involvement

Patients were not involved in design, development of research question and study tools and recruitment of study participants.

## RESULTS

This cross-sectional study was carried out in a tertiary care hospital with a sample size of 180. This included 80 antenatal mothers, 80 postnatal mothers and 20 staff nurses.

Step 1 is about having a breastfeeding policy. Written breastfeeding policy (step 1) showed full compliance as IEC banners and posters were displayed in wards and consultation rooms. All the antenatal, postnatal and labor wards displayed written breastfeeding policy (100%) and there are no posters or materials promoting breast milk substitutes (100%). Step 2 is about training the health care workers about the breastfeeding policy. Out of the 20 staffs in the study, 4 staffs received 18 hours of training according to records and 4 staffs received 18 hours of training as reported by them, yet all the 20 have the correct breastfeeding management knowledge. There is not any staff scheduled for refresher training. (Table 1)

**Table 1:** Compliance of baby friendly hospital initiative (Steps 1-5)

| Step 1– have a breastfeeding policy that is routinely communicated to all healthcare staff |  |  |  |  |
| --- | --- | --- | --- | --- |
| Step1 | Criteria |  |  | Percentage |
| 1(A) | Policy displayed |  |  | 100% |
| 1(B) | No posters or materials promoting breast milk substitutes |  |  | 100% |
| Cumulative mean % for step 1 [(1a + 1b)/2] = 100% |  |  |  |  |
| Step 2– Train all health care staff in skills necessary to implement this policy |  |  |  |  |
| Step2 | Criteria |  | N=20 | Percentage |
| 2(A) | Staff receiving 18 hours training (In records) |  | 4 | 20 |
| 2(B) | Staff receiving 18 hours training (reported by staff) |  | 4 | 20 |
| 2(C) | Staff with correct breastfeeding management knowledge |  | 20 | 100 |
| 2(D) | Trained staff scheduled for refresher training |  | 0 | 0 |
| Cumulative mean % for step 2 [(2a + 2b + 2c + 2d)/4] = 35% |  |  |  |  |
| Step 3–inform all pregnant women about benefits and management of breastfeeding |  |  |  |  |
| 3(A) | 1 | Written description of prenatal education |  | 100% |
| 3(b) | Topic | Knowledge of mothers on: | N = 80 | Percentage |
|  | 1 | Benefits of breast feeding | 55 | 68.75 |
|  | 2 | Importance of breastfeeding soon after delivery | 60 | 75 |
|  | 3 | Importance of rooming in | 29 | 36.25 |
|  | 4 | Correct positioning and attachment | 38 | 47.5 |
|  | 5 | Importance of feeding on demand | 50 | 62.5 |
|  | 6 | What can be done to ensure production of enough breastmilk | 42 | 52.5 |
|  | 7 | Importance of giving only breastmilk up to 6 months | 58 | 72.5 |
|  |  | Mean |  | 59.29% |
| Cumulative mean % for step 3 [(3a + 3b)/2] = 79.64% |  |  |  |  |
| Step 4: Help mothers initiate breast feeding within half-hour of birth |  |  |  |  |
| 4(a) | Babies breastfed within one hour of birth (vaginal delivery) |  | 25 (Out of 43) | 58.13% |
| 4(b) | Babies breastfed within one hour of mothers being able to respond (caesarean) |  | 16 (Out of 37) | 43.24% |
| 4(c) | Babies breastfed within one hour (vaginal and caesarean) |  | 41 (Out of 80) | 51.25% |
| Cumulative mean % for step 4 [(4a + 4b)/2] = 51.25% |  |  |  |  |

Step 3 is about educating the pregnant women about benefits and management of breastfeeding. Maternal and child protection (MCP) card was available with all the antenatal mothers, which contained information regarding breastfeeding practices. Also, all discharge summaries of antenatal mothers admitted during their pregnancy had details of breastfeeding. Written information about breastfeeding was given to all prenatal mothers (100%) yet, only 75% knew the importance of breastfeeding soon after delivery and 72.2% knew the importance of giving only breast milk up to 6 months. 68.75% of mothers knew the benefits of breastfeeding, 62.5% knew the importance of feeding on demand and 52.5% knew how to ensure enough production of breast milk. Only 47.5% of prenatal mothers had knowledge on correct positioning and attachment of the baby and 36.25% knew about rooming in.

51.3% of the babies were breastfed within one hour of birth (Step 4) and the proportion was even lower (43%) for infants delivered via caserean section. (Table 1)

50% of the mothers were offered help with breastfeeding(Step 5).63% of the mothers were offered help with correct positioning or attachment.63% of the mothers demonstrated correct positioning/attachment and 61% of the mothers were taught how to express milk. Only 20% of the staff had demonstrated correct positioning/attachment though majority of them said that they knew the correct technique. Step 6 is regarding exclusive breastfeeding and 62.5% of babies were exclusively breastfed. (Table 2).

**Table 2:**
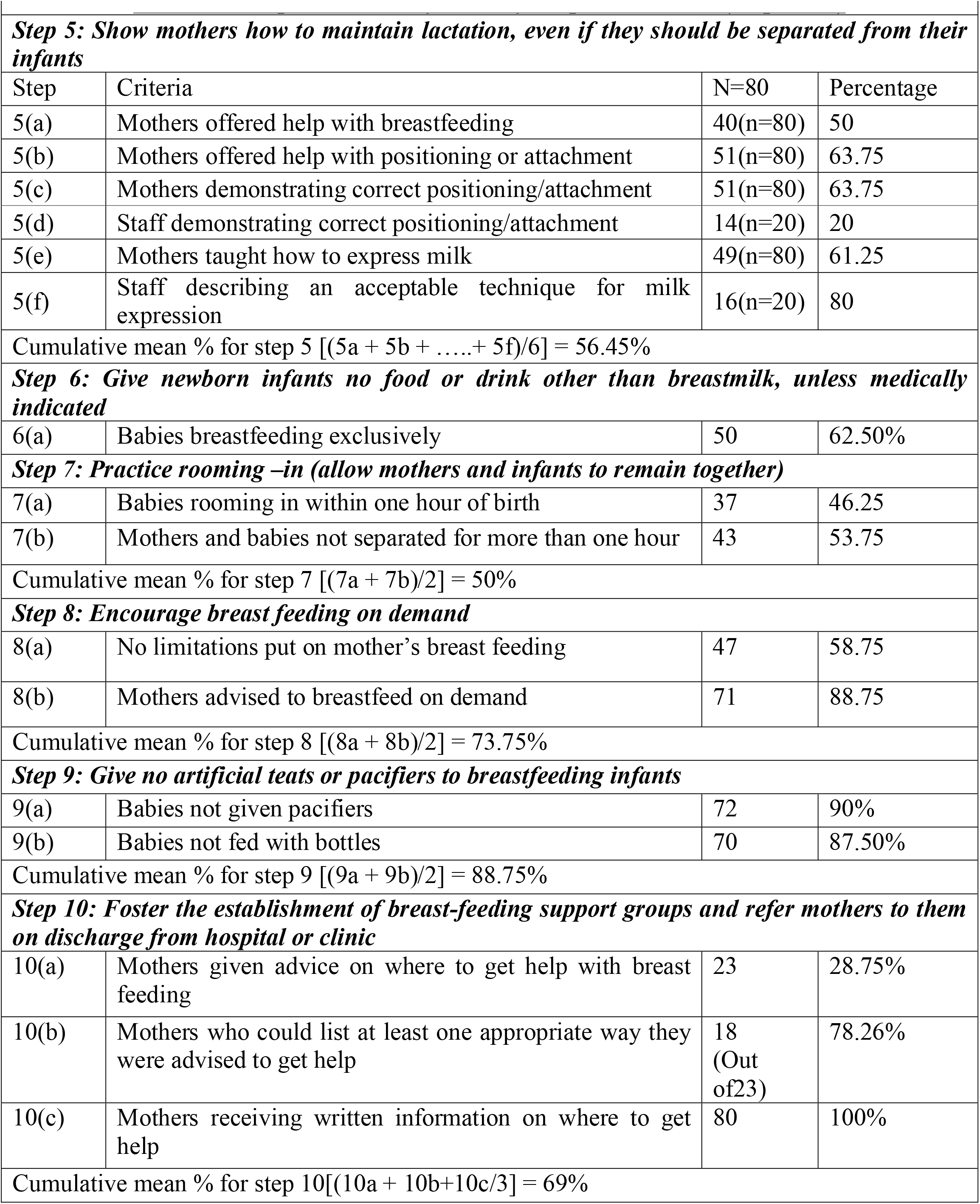
Compliance of baby friendly hospital initiative (Steps 6-10)

Step 7 is about rooming in and 46.25% of postnatal mothers practiced rooming in within one hour of birth.53.75% of mothers were not separated from their babies for more than one hour. Mean score of steps 7 was 50% (Table 2).

Step 8 is about encouraging breast feeding on demand.88.75% of postnatal mothers were advised to breast feed on demand and no limitations were put on mothers in their breast feeding in 58.75%. Mean score of steps 8 was 73.75% (Table 2).

Step 9 is about not giving any artificial teats or pacifiers to breastfeeding infants.90% were not given pacifiers and 87.5% were not fed with bottle. Mean score of steps 9 was 88.75% (Table 2).

Step 10 is about breast-feeding support groups. 100% of mothers were given written information on where to get help with breast feeding after discharge, while only 28.5% mothers were given advice regarding the same. Out of the 23 mothers who were given advice on where to get help with breast feeding, 78.26% of them could list at least one appropriate way they were advised to get help. Mean score of steps 10 was 88.75% (Table 2).

Step 1 (written breast-feeding policy) was fully met, hence has good compliance. Step 2 (staff training) was least met, thus resulting in poor compliance. The rest of the steps were moderately met.

Compliance with the Ten Steps of baby friendly initiative was 66.63%, signifying a moderate compliance overall.

## DISCUSSION

This cross-sectional study was carried out in a tertiary care hospital with a sample size of 180. This included 80 antenatal mothers, 80 postnatal mothers and 20 staff nurses.

Written breastfeeding policy (step 1) showed full compliance. Only 35% of the staff nurses interviewed received formal training in breastfeeding techniques which is comparable to national trends.^[14]^ Of all steps, step 2 (train all healthcare staff) has the least compliance due to there being a yearly rotation of staff in various departments, high staff turnover and funding constraints. The 2018 revision of the BFHI criteria focuses on practical competencies (WHO & UNICEF, 2018) necessitating flexible and less time-consuming training.^[5]^ Available evidences suggest that implementation of minimum of 18 hours training for health care providers has been shown to positively impact the breast feeding practices significantly.^[14]^ Though concrete efforts are taken by the government to promote breastfeeding, proportion of trained staff nurses availability at labour wards is still low.^[7]^ TS Raghu Raman et al has demonstrated that proper health care providers’ education and training can lead to the modification of practises from being “inappropriately baby friendly” to being “appropriately baby and mother friendly.^[15]^

Only 51% of the babies were breastfed within one hour and it is lower than the national average of 55% ^[16]^which is a worrying trend. Practice of breastfeeding within one hour reported by us is low as compared to other studies as well. Sridevi et al had reported that 59% of mothers initiated breastfeeding within 1 h of childbirth.^[17]^ Shalini S et al. has reported that 56% of mothers initiated breastfeeding within 1 h of childbirth.^[18]^ The national family health survey 5 reports that 60% of mothers had initiated breastfeeding within one hour in Tamil Nadu.^[19]^ Training component and monitoring mechanisms needs to be strengthened more to sustain the good breast feeding practices.^[16]^

75% of the post-natal mothers knew the importance of breastfeeding soon after delivery and 72.2% knew the importance of giving only breast milk upto 6 months. Only 47.5% of prenatal mothers had knowledge on correct positioning and attachment of the baby and 36.25% knew about rooming in. Jelly et all had reported better results than our study(81%).^[20]^

Step 5 (how mothers breastfeed/ maintain lactation) was moderately followed (56.45%) (Table 2). Only 50 % of them were offered help with breastfeeding, 63.75% of them were offered help with positioning and attachment of the baby and hence the same 63.75% of them were able to demonstrate correct method of positioning and attachment. Sultania et al had reported good compliance of breastfeeding techniques^[21]^ whereas some studies had reported low compliance of good breastfeeding techniques.^[22]^

Only 62.5% of the babies were exclusively breastfed in our studies which is above the national family health survey-5 finding of 55%.^[19]^However, few studies had reported higher percentage.^[18]^This area didn’t show much progress over the period of years though there is strong evidence that exclusive breastfeeding is essential up to 6 months of age of the infant. Breastfeeding is a complex behavior and mere knowledge is not enough to sustain the practice. Lot of factors influence the compliance of breastfeeding like cultural beliefs, strong influence from the commercial sector, lack of support and mis information etc.^[9]^

Despite there being a written breastfeeding policy on the maternal child health card (MCH), there is moderate compliance to step 3 (inform all pregnant women on breastfeeding) and step 8 (encourage breastfeeding on demand) because mothers often tend to skip reading that section of the card. Other reasons for poor adherence to step 8 were sore nipples, unsupportive households, working mothers and so on.

According to Gupta et al., Steps 3, 5, and 10 that call for expertise and depend on the education and availability of healthcare professionals appear to be more challenging to implement.^[14]^ The overall compliance of the institution under study, with the BFHI standards was found to be moderate viz, 66.63%. In a study done in Ganna, the compliance was 86%.^[10]^According to Cochrane review on baby friendly hospital initiative, average global BFHI compliance score is 77 which is higher than our study compliance score.^[23]^ According to World bank report, though the hospital staff was required to promote early breastfeeding and encourage mothers who wanted to breastfeed exclusively, these measures were not supported by any policy, monitoring system, or specialised training. Particularly during caesarean section deliveries, newborns were separated from their mothers. Mothers with breastfeeding problems lacked any sort of follow-up support from the healthcare system.^[24]^

## CONCLUSION

The compliance to Ten steps of BFHI in low resource setting shows an overall moderate compliance. To attain the intended results, evaluation tool development for assessing breastfeeding habits and rewarding healthcare organizations should be promoted. The Ten Steps of BFHI may be implemented more effectively in all medical facilities through the Mother’s absolute affection Program(MAA).

More focused approaches are needed to improve the breastfeeding practices even in tertiary care settings in India.

## LIMITATIONS

Staff could have altered their behaviors and given responses that might not necessarily reflect their routine practices.

The study was conducted in a tertiary hospital in the government sector. Therefore, its findings and conclusions are not applicable to the primary and secondary health facilities in the government sector or to the private hospitals.

The study was conducted at a single centre and therefore its results cannot be accurately generalized on a large scale.

## Data Availability

All data produced in the present study are available upon reasonable request to the authors

## REFERENCES

1. Victora CG, Bahl R, Barros AJD, França GVA, Horton S, Krasevec J, et al. Breastfeeding in the 21st century: epidemiology, mechanisms, and lifelong effect. Lancet Lond Engl. 2016 Jan 30;387(10017):475–90.

2. Weltgesundheitsorganisation, UNICEF, editors. Global strategy for infant and young child feeding. Geneva: WHO; 2003. 30 p.

3. Smith ER, Hurt L, Chowdhury R, Sinha B, Fawzi W, Edmond KM, et al. Delayed breastfeeding initiation and infant survival: A systematic review and meta-analysis. PloS One. 2017;12(7):e0180722.

4. Protecting, promoting and supporting breastfeeding: the Baby-friendly Hospital Initiative for small, sick and preterm newborns. Geneva: World Health Organization and the United Nations Children’s Fund (UNICEF), 2020.

5. World Health Organization, United Nations Children’s Fund (UNICEF). Implementation guidance: protecting, promoting and supporting breastfeeding in facilities providing maternity and newborn services: the revised baby-friendly hospital initiative [Internet]. Geneva: World Health Organization; 2018 [cited 2023 Jan 16]. Available from: https://apps.who.int/iris/handle/10665/272943

6. Hofvander Y. Breastfeeding and the Baby Friendly Hospitals Initiative (BFHI): organization, response and outcome in Sweden and other countries. Acta Paediatr Oslo Nor 1992. 2005 Aug;94(8):1012–6.

7. MAA-Mother’s absolute affection:Programme for promotion of breast feeding, Operational guidelines,MOHFW,GOI, 2016 [Internet]. [cited 2023 Jan 17]. Available from: https://nhm.gov.in/MAA/Operational_Guidelines.pdf

8. ‘Arrested Development; 5 th Assessment of India’s Policy & Programmes on Infant & Young Child Feeding,Breastfeeding Promotion Network of India (BPNI)/IBFAN Asia 2018 [Internet]. [cited 2023 Jan 16]. Available from: https://www.worldbreastfeedingtrends.org/uploads/country-data/country-report/WBTi-India-Report-2018.pdf

9. Jp Dadhich, Gupta A, Prasad V, Bhar RH, Gaur A, Anurag Singh. WBTi India Assessment Report 2008. 2008 [cited 2023 Jan 16]; Available from: http://rgdoi.net/10.13140/RG.2.2.20081.99683

10. Agbozo F, Ocansey D, Atitto P, Jahn A. Compliance of a Baby-Friendly Designated Hospital in Ghana With the WHO/UNICEF Baby and Mother-Friendly Care Practices. J Hum Lact Off J Int Lact Consult Assoc. 2020 Feb;36(1):175–86.

11. Competency verification toolkit: ensuring competency of direct care providers to implement the Baby-friendly Hospital Initiative. Geneva: World Health Organization and the United Nations Children’s Fund (UNICEF), 2020.

12. World Health Organization, Wellstart International & United Nations Children’s Fund (UNICEF). (1999). The baby-friendly hospital initiativeLJ: monitoring and reassessmentLJ: tools to sustain progress-section II / prepared by the World Health Organization and Wellstart International. World Health Organization. https://apps.who.int/iris/handle/10665/65380 [Internet]. [cited 2023 Jan 17]. Available from: https://apps.who.int/iris/bitstream/handle/10665/65380/WHO_NHD_99.2_%28sectionI-II%29.pdf?sequence=1&isAllowed=y

13. World Health Organization, Wellstart International & United Nations Children’s Fund (UNICEF). (1999). The baby-friendly hospital initiativeLJ: monitoring and reassessmentLJ: tools to sustain progress-Section III-IV / prepared by the World Health Organization and Wellstart International. World Health Organization. https://apps.who.int/iris/handle/10665/65380 [Internet]. [cited 2023 Jan 17]. Available from: https://apps.who.int/iris/bitstream/handle/10665/65380/WHO_NHD_99.2_%28sectionIII-IV%29.pdf?sequence=2&isAllowed=y

14. Gupta DA. Report of All India Study-Newborn Care, Infant Feeding Practices and Implementation of the “Infant Milk Substitutes, Infant Foods and Feeding Bottles (Regulation of Production, Distribution and Supply) Act, 1992.” in the Hospital Settings, and Infant Feeding Practices in the Catchment Area of These Hospitals.

15. Raman TR, Parimala V, Iyengar A. Baby friendly hospital initiative experiences from a service hospital. Med J Armed Forces India. 2001 Jan;57(1):22–5.

16. South-Asia-Baby-Friendly-Hospital-Initiative-in-South-Asia-Implementing-Ten-Steps-to-Successful-Breastfeeding-India-Nepal-and-Bangladesh-Challenges-and-Opportunities.pdf [Internet]. [cited 2023 Feb 6]. Available from: https://documents1.worldbank.org/curated/en/916891573111241173/pdf/South-Asia-Baby-Friendly-Hospital-Initiative-in-South-Asia-Implementing-Ten-Steps-to-Successful-Breastfeeding-India-Nepal-and-Bangladesh-Challenges-and-Opportunities.pdf

17. Naaraayan S, Priyadharishini D, Geetha R, Vengatesan A. SOCIOECONOMIC DETERMINANTS OF BREASTFEEDING PRACTICES IN SOUTH INDIA - A HOSPITAL-BASED CROSS-SECTIONAL STUDY. Indian J Child Health. 2018 Jan 25;05:50–3.

18. S S, S G. Breastfeeding practices of nursing mothers in Tamil Nadu: a hospital based cross sectional study. Int J Community Med Public Health. 2018 Sep 24;5(10):4441–9.

19. National Family Health Survey 2019-21,Tamil Nadu factsheet,MOHFW,GOI [Internet]. [cited 2023 Jan 17]. Available from: http://rchiips.org/nfhs/NFHS-5_FCTS/Tamil_Nadu.pdf

20. Jelly P, Kodi M, Sharma M, Sharma SK, Sharma R. Knowledge, preferences, practices, and attitudes about breastfeeding among postnatal mothers in Uttarakhand, India: a cross-sectional study. Indian J Community Health. 2022 Jun 30;34(2):294–300.

21. Sultania P, Agrawal NR, Rani A, Dharel D, Charles R, Dudani R. Breastfeeding Knowledge and Behavior Among Women Visiting a Tertiary Care Center in India: A Cross-Sectional Survey. Ann Glob Health. 85(1):64.

22. Koya S, Babu GR R D, Iyer V, Yamuna A, Lobo E, et al. Determinants of Breastfeeding Practices and Its Association With Infant Anthropometry: Results From a Prospective Cohort Study in South India. Front Public Health [Internet]. 2020 [cited 2023 Jan 17];8. Available from: https://www.frontiersin.org/articles/10.3389/fpubh.2020.492596

23. Maastrup R, Haiek LN, Group TN-BS. Compliance with the “Baby-friendly Hospital Initiative for Neonatal Wards” in 36 countries. Matern Child Nutr. 2019;15(2):e12690.

24. World Bank. 2019. Baby-friendly Hospital Initiative (BFHI) in South Asia: Implementing Ten Steps to Successful Breastfeeding. India, Nepal and Bangladesh, Challenges and Opportunities. © World Bank [Internet]. [cited 2023 Feb 6]. Available from: https://documents1.worldbank.org/curated/en/916891573111241173/pdf/South-Asia-Baby-Friendly-Hospital-Initiative-in-South-Asia-Implementing-Ten-Steps-to-Successful-Breastfeeding-India-Nepal-and-Bangladesh-Challenges-and-Opportunities.pdf

